# Causal associations between COVID-19 and Atrial Fibrillation: A bidirectional Mendelian randomization study

**DOI:** 10.1101/2020.08.13.20174417

**Authors:** Xiaoyu Zhang, Biyan Wang, Di Liu, Jinxia Zhang, Qiuyue Tian, Xiaoni Meng, Jie Zhang, Haifeng Hou, Manshu Song, Wei Wang, Youxin Wang

**Author notes:** Xiaoyu Zhang and Biyan Wang contributed equally to this paper and should share the first authorship. **Correspondence**: Youxin Wang, PhD, Professor, School of Public Health, Capital Medical University, No.10 Xitoutiao, Youanmenwai Street, Fengtai District, Beijing, 100069, China.

## Abstract

**Background:** Observational studies showed that coronavirus disease 2019 (COVID-19) attacks universally and its most menacing progression uniquely endangers the elderly with cardiovascular disease (CVD). Whether COVID-19 is causally related to increasing susceptibility and severity of atrial fibrillation (AF), the main form of CVD, remains still unknown.

**Methods:** The study aims to investigate the bidirectional causal relations of COVID-19 with AF using two-sample Mendelian randomization (MR) analysis.

**Results:** MR evidence suggested genetically predicted severe COVID-19 was significantly associated with higher risk of AF (odds ratio [OR], 1.041; 95% confidence interval (CI), 1.007-1.076; *P* = 0.017), while genetically predicted AF was not causally associated with severe COVID-19 (OR, 0.831; 95% CI, 1.619-1.115; *P*=0.217). There was limited evidence to support association of genetically proxied COVID-19 with risk of AF (OR, 1.051; 95% CI, 0.991-1.114; *P*=0.097), and vice versa (OR, 0.163; 95% CI, 0.004-6.790; *P*=0.341). MR-Egger indicated no evidence of pleiotropic bias.

**Conclusion:** Overall, severe COVID-19 may causally affect AF through independent biological pathway. Survivors from severe COVID-19 might be of high risk of AF in the future.

## Introduction

Coronavirus disease (COVID-19), which is caused by the severe acute respiratory syndrome coronavirus 2 (SARS-COV2) and represents the causative agent of a potentially fatal disease, rapidly emerged as a global pandemic and afflicted global finances and healthcare systems severely^1^. The virus attacks universally and is vulnerable to the elderly, especially those with cardiovascular comorbidities such as diabetes mellitus, hypertension, heart failure, and coronary heart disease^2^. COVID-19 may cause adverse impact on the heart and cardiovascular system. Recent studies have found that SARS-COV2 infection may damage cardiac myocytes and increase atrial fibrillation (AF) risk^3-5^. However, those findings are still susceptible to confounding and reverse causation that cannot be fully ruled out in observational studies.

Mendelian randomization (MR) is a burgeoning field that utilizes genetic variants that are robustly associated with such modifiable exposures to generate more reliable evidence^6^. This approach relies on the natural, random assortment of genetic variants during meiosis yielding a random distribution of genetic variants^7^. Genome-wide association studies (GWAS) data, which typically provide regression coefficients summarizing the associations of many genetic variants with various traits, are potentially a powerful source of data for MR analysis^8^. Therefore, we performed bidirectional MR analyses for causal inference between COVID-19 and AF using summary statistics GWAS results of COVID-19 and AF. Understanding the bidirectional relations between COVID-19 and AF is of significant public health importance about disease prevention and management of complications.

## Methods

### Data sources

#### Genetic association datasets for COVID-19

Summary genetic association estimates for risk of COVID-19 were obtained from the most recent version of GWAS analyses of the COVID-19 host genetics initiative in UK Biobank individuals released on July 1, 2020 (https://www.covid19hg.org/results/)^9^. We selected two phenotypes from this GWAS: (1) Covid *vs*. lab/self-reported negative including 3,523 patients and 36,634 control participants; (2) very severe respiratory confirmed Covid *vs*. population including 536 patients and 329,391 control participants.

In addition, summary genetic association estimates for risk of severe COVID-19 with respiratory failure were reported by Severe COVID-19 GWAS Group including 1610 cases and 2205 controls in Italy and Spain^10^.

#### Genetic association datasets for AF

We drew on summary statistics from a recent GWAS of AF which was conducted by FinnGen Study including 12,859 patients and 73,341 control participants https://www.finngen.fi/en). All GWAS summary statistic data were based on European ancestry population.

### Instrumental variables for COVID-19 and AF

We first performed forward MR analysis to assess the effect of the phenotypes of COVID-19 or severe COVID-19 on AF by using genetic variants associated with COVID-19 or severe COVID-19 as instrumental variables (IVs). Single nucleotide polymorphisms (SNPs) were selected as IVs for COVID-19 or severe COVID-19 associated at *P*<1×10^-5^, because we didn’t filter out SNPs based on *P*<5×10^-8^ from the COVID-19 host genetics initiative. In addition, we chose two SNPs (rs11385942 and rs657152) as IVs for severe COVID-19 from Severe Covid-19 GWAS Group based on *P*<5×10^-8^. However, for rs657152 that was not present in FinnGen Study, we failed to find proxy.

Then, we conducted reverse MR using genetic variants associated with AF as IVs to investigate their effect on COVID-19 and severe COVID-19. SNPs that achieved significance (*P*<5×10^-8^) for the AF were selected as IVs.

We only retained independent variants from each other based on European ancestry reference data from the 1000 Genomes Project (Linkage disequilibrium [LD], r^2^ threshold = 0.0001).

### MR analysis

In the main analyses, we combined MR estimates for each direction of potential influence using inverse variance-weighted (IVW) meta-analysis, which actually was a weighted regression of SNP-outcome effects on SNP-exposure effects (the intercept is constrained to 0)^8^. Results can be biased if IVs show horizontal pleiotropy, affecting the outcome through other pathways other than the exposure which could violate MR assumptions^11^. Therefore, we performed sensitivity analyses including the weighted median (WM), penalised weighted median (PWM), Pleiotropy Residual Sum and Outlier (MR-PRESSO) and MR-Egger regression, which are known to be relatively robust to horizontal pleiotropy and invalid instruments. The WM method which selects the median MR estimate as the causal estimate may provide precise causal estimates against invalid instruments^12^. MR-PRESSO was applied to detect and correct for any outliers reflecting likely pleiotropic biases for all reported results^13^. We conducted MR-Egger analysis which allows the intercept to be freely assessed as an indicator of average pleiotropic bias^11^. In order to assess robustness of significant results, we applied further tests for horizontal pleiotropy to detect heterogeneous outcomes, including leave one out analysis, the Cochran Q statistic, and the MR Egger intercept test of deviation from the null^14^. Results are presented as odds ratios (ORs) with 95% confidence intervals (95% CIs) of outcomes per genetically predicted increase in each exposure factor.

For bidirectional MR analyses, the causal relationships between COVID-19 and AF were delineated into four potential parts. **Figure 1** showed an overview of the bidirectional MR study to investigate these explanations. If the *P* value was less than 0.05 only in forward MR for Explanation 1, there was significant association of genetically instrumented COVID-19 with higher AF risk. Then we conducted the reverse MR analysis assessing whether AF affected COVID-19. This reverse causal association was existed if *P* value was less than 0.05 in Explanation 2. The Explanation 3 showed there were bidirectionally causal associations between COVID-19 and AF (*P* <0.05). There was no any causal association in forward and reverse MR (*P* >0.05) as showed in Explanation 4.

**Figure. 1.**
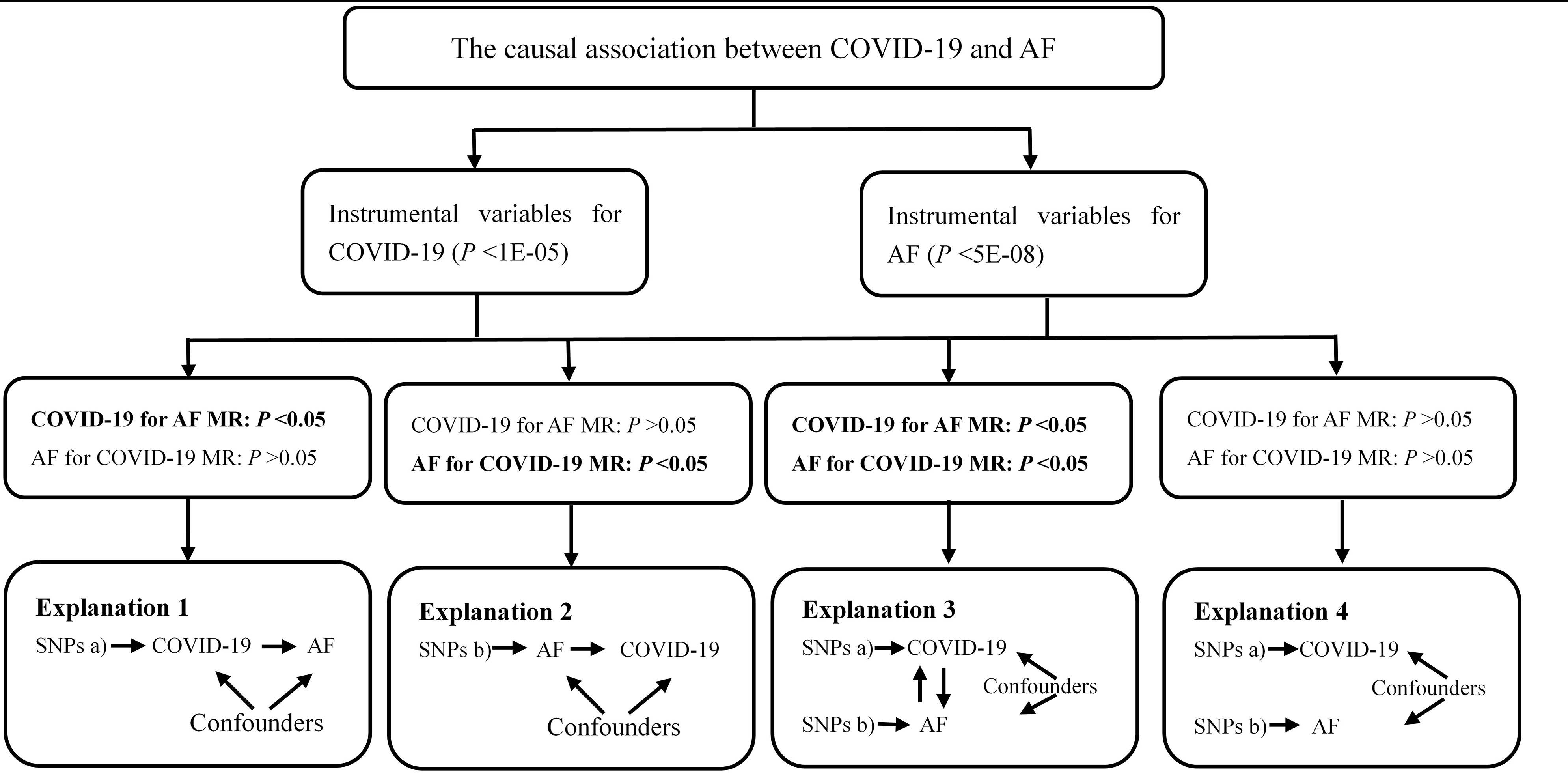
Analyses pipeline to evaluate the explanations for the observed associations between COVID-19 and AF COVID-19: Coronavirus disease 2019; AF: Atrial fibrillation; MR: mendelian randomization; SNP, single nucleotide polymorphism

All data analyses for MR were conducted by “twosampleMR” and “MR-PRESSO” packages in the R environment (R version 3.6.3, R Project for Statistical Computing). This package harmonizes exposure and outcome data sets including information on SNPs, alleles, effect sizes, standard errors, *P* values, and effect allele frequencies for selected exposure instruments.

### Results

#### Causal association of COVID-19 with AF via forward MR

The summary genetic associations data were reported in the Supplement Table 1. In the forward MR analysis, we used 16 independent SNPs as the IVs for COVID-19. As shown in Table 1, IVW estimate showed there was no association between the genetically instrumented COVID-19 and AF risk (OR, 1.051; 95% CI, 0.991-1.114; *P*=0.097), without notable heterogeneity (*P*=0.782) across instrument SNP effects. The MR Egger intercept test further suggested no horizontal pleiotropy (intercept= -0.004, *P*=0.842). Of note, there was association of the genetically instrumented severe COVID-19 with AF using 13 SNPs (OR, 1.041; 95% CI, 1.007-1.076, *P*=0.017, without directional pleiotropy (intercept=0.034, *P*=0.205) and heterogeneity (*P*=0.371).

**Table 1.** Bidirectional causal relations of COVID-19 with AF performed by MR analyses

| *Causal association of COVID-19 with AF via forward MR* |  |  |  |  |
| --- | --- | --- | --- | --- |
| Phenotype | Numbers of SNPs | OR (95% CI) | Beta (SE) | *P* |
| COVID vs. Lab/self-reported negative |  |  |  |  |
| IVW | 16 | 1.051 (0.991-1.114) | 0.049 (0.030) | 0.097 |
| Weighted median | 16 | 1.054 (0.971-1.143) | 0.052 (0.042) | 0.208 |
| Penalised weighted median | 16 | 1.054 (0.968-1.147) | 0.052 (0.043) | 0.228 |
| MR-PRESSO | 16 | 1.068 (1.015-1.121) | 0.066 (0.027) | 0.025 |
| MR-Egger | 16 | 1.064 (0.931-1.215) | 0.062 (0.068) | 0.379 |
| $\beta$ (intercept) | - | - | -0.004 (0.019) | 0.842 |
| Q statistic | - | - | - | 0.782 |
| Severe respiratory confirmed COVID vs. population |  |  |  |  |
| IVW | 13 | 1.041 (1.007-1.076) | 0.040 (0.017) | 0.017 |
| Weighted median | 13 | 1.033 (0.987-1.081) | 0.032 (0.023) | 0.163 |
| Penalised weighted median | 13 | 1.033 (0.987-1.081) | 0.032 (0.023) | 0.158 |
| MR-PRESSO | 13 | 1.036 (1.006-1.065) | 0.035 (0.015) | 0.033 |
| MR-Egger | 13 | 0.964 (0.858-1.083) | -0.369 (0.060) | 0.548 |
| $\beta$ (intercept) | - | - | 0.034 (0.025) | 0.205 |
| Q statistic | - | - | - | 0.371 |
| *Causal association of AF with COVID-19 via reverse MR* |  |  |  |  |
| COVID vs. Lab/self-reported negative |  |  |  |  |
| IVW | 6 | 0.163 (0.004-6.790) | -1.812 (1.902) | 0.341 |
| Weighted median | 6 | 0.126 (-) | -2.073 (15.556) | 0.894 |
| Penalised weighted median | 6 | 0.126 (00003-5.682) | -2.073 (3.119) | 0.506 |
| MR-PRESSO | 6 | 0.901 (0.848-0.954) | -0.104 (0.068) | 0.163 |
| MR-Egger | 6 | 0.007 (0.00002-2.580) | -4.957 (3.013) | 0.175 |
| $\beta$ (intercept) | - | - | -0.0007 (0.0028) | 0.265 |
| Q statistic | - | - | - | 2.565e-97 |
| Severe respiratory confirmed COVID vs. population |  |  |  |  |
| IVW | 8 | 0.831 (0.619-1.115) | -0.186 (0.150) | 0.217 |
| Weighted median | 8 | 0.826 (0.579-1.178) | -0.191 (0.181) | 0.292 |
| Penalised weighted median | 8 | 0.826 (0.578-1.181) | -0.191 (0.183) | 0.295 |
| MR-PRESSO | 8 | 0.830 (0.677-0.983) | -0.186 (0.078) | 0.049 |
| MR-Egger | 8 | 0.726 (0.409-1.287) | -0.321 (0.292) | 0.315 |
| $\beta$ (intercept) | - | - | 0.033 (0.063) | 0.610 |
| Q statistic | - | - | - | 0.966 |

Beta is the estimated effect size. $P < 0.05$ were considered statistically significant.
AF: Atrial Fibrillation; CI: confidence intervals; IVs: instrumental variables; IVW, inverse-variance weighted; MR mendelian randomization;
MR-PRESSO: Pleiotropy Residual Sum and Outlier; OR: odds ratio; SE, standard error; SNP, single-nucleotide polymorphism.

In sensitivity analyses, nominally significant associations of genetically instrumented COVID-19 (OR, 1.068; 95% CI, 1.015-1.121, *P*=0.025) and severe COVID-19 (OR, 1.036; 95% CI, 1.006-1.065, *P*=0.033) were observed for the risk of AF using MR-PRESSO methods. The results of leave-one-out sensitivity analyses showed that the causal associations between genetically instrumented COVID-19 and severe COVID-19 and AF was not substantially driven by any individual SNP (Figure 2 A and 2B). For IVs, MR-PRESSO did not detect any potential outliers. Associations between each variant with severe COVID-19 and risk of AF was displayed in Figure 3.

**Figure. 2A.**
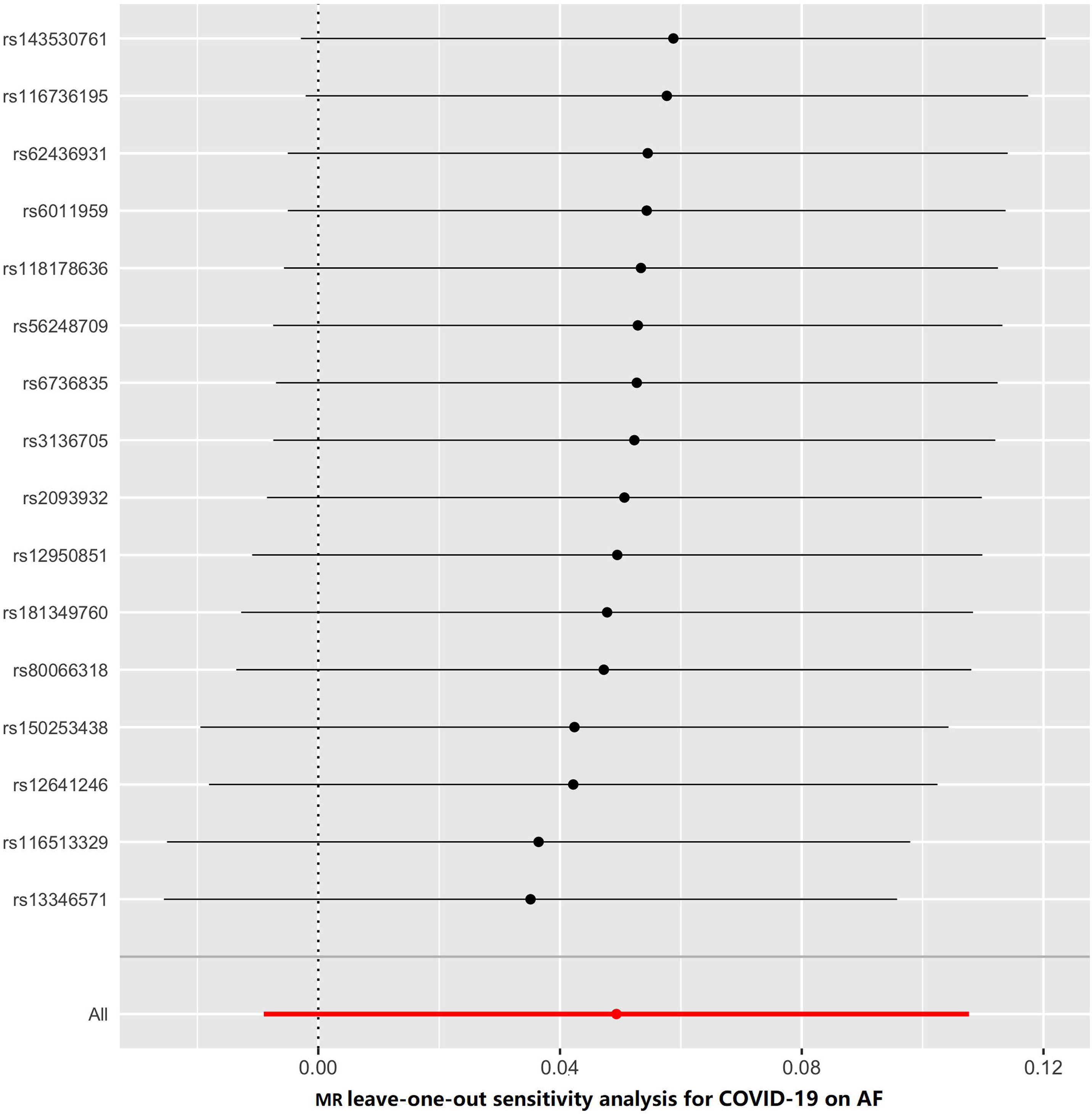
MR leave-one-out analysis for COVID-19 on AF. Each row represents a MR analysis of COVID-19 on AF using all instruments expect for the SNP listed on the y-axis. The point represents the odds ratio with that SNP removed and the line represents 95% confidence interval. COVID-19: Coronavirus disease 2019; AF: Atrial fibrillation; MR: mendelian randomization; SNP, single nucleotide polymorphism

**Figure. 2B.**
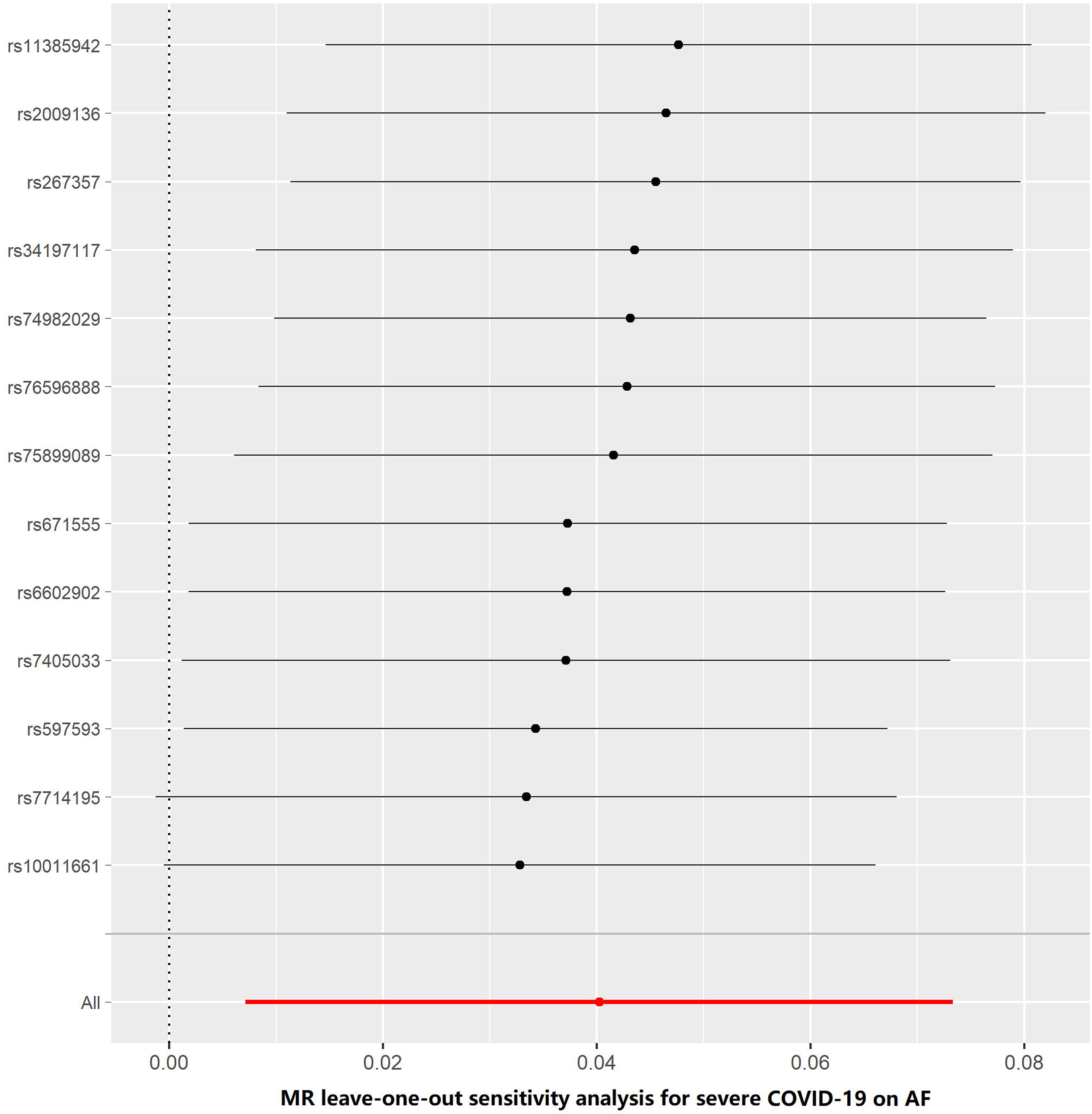
MR leave-one-out analysis for severe COVID-19 on AF. Each row represents a MR analysis of using all instruments expect for the SNP listed on the y-axis. The point represents the odds ratio with that SNP removed and the line represents 95% confidence interval. COVID-19: Coronavirus disease 2019; AF: Atrial fibrillation; MR: mendelian randomization; SNP, single nucleotide polymorphism

#### Causal association of AF with COVID-19 via reverse MR

The summary genetic associations data of AF were reported in the Supplement Table 2. As shown in Table 1, the reverse MR analysis showed no significant association of genetically instrumented AF with COVID-19 using 6-SNP (OR, 0.163; 95% CI, 0. 004-6.790; *P*=0.341). Pleiotropy bias (intercept= -0.0007, *P*=0.265) was not detected and heterogeneity (*P*=2.565e-97) was found. Moreover, we found no statistically significant evidence of a relationship between AF and severe COVID-19 using 8-SNP (OR, 0.831; 95% CI, 1.619-1.115; *P*=0.217). There was no heterogeneity and horizontal pleiotropy based on the Q test (*P*=0.966) and MR-Egger intercept test (*P*=0.610). MR-PRESSO did not detect any potential outliers for IVs. The similar results were found in the sensitivity analyses. The results of leave-one-out sensitivity analysis showed that the association between genetically instrumented AF with severe

COVID-19 was not substantially driven by any individual SNP (Figure 2D), while the association between genetically instrumented AF with COVID-19 was driven by rs17042121 (Figure 2C).

**Figure. 2C.**
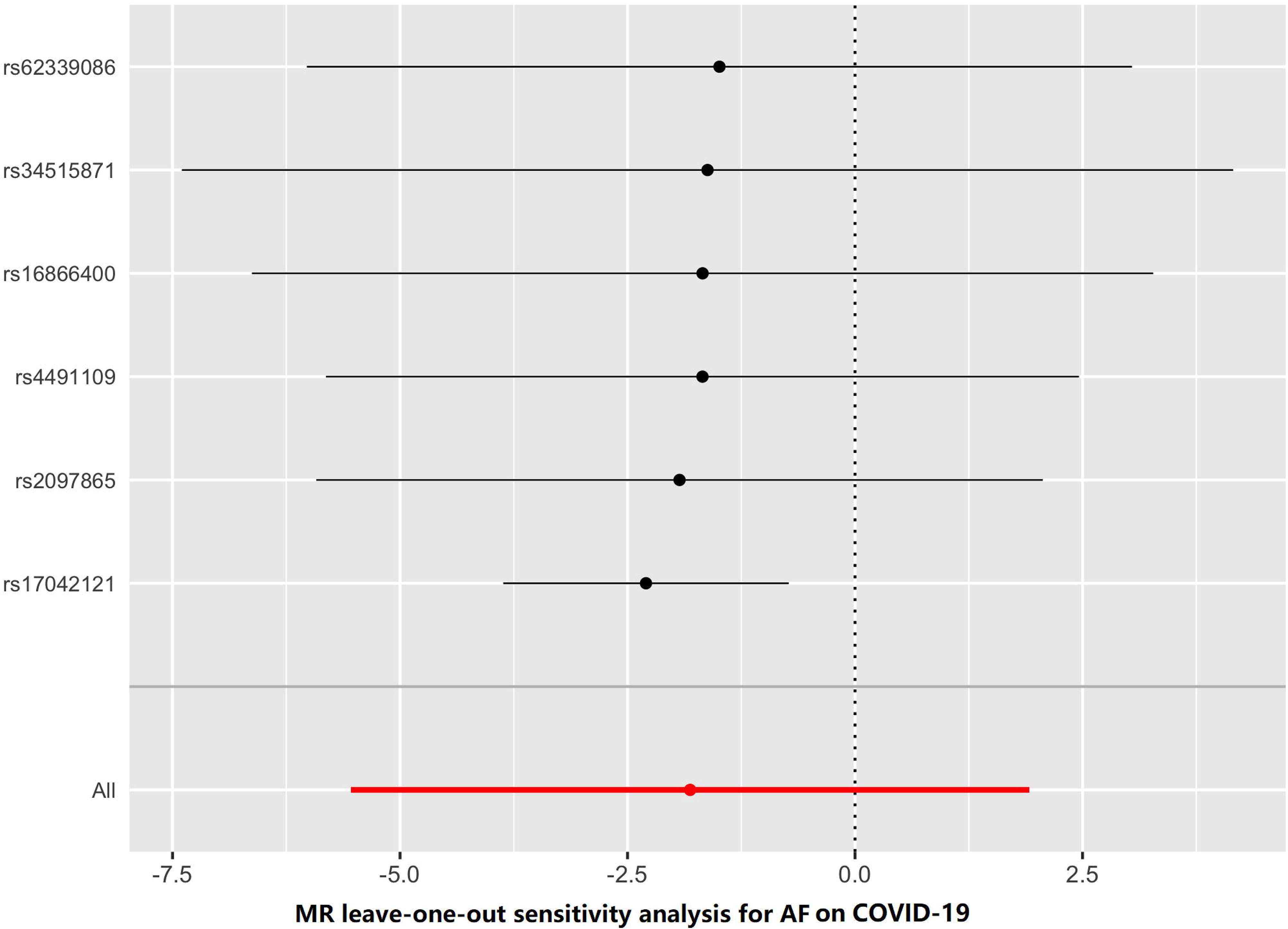
MR leave-one-out analysis for AF on COVID-19. Each row represents a MR analysis of AF on COVID-19 using all instruments expect for the SNP listed on the y-axis. The point represents the odds ratio with that SNP removed and the line represents 95% confidence interval. COVID-19: Coronavirus disease 2019; AF: Atrial fibrillation; MR: mendelian randomization; SNP, single nucleotide polymorphism

**Figure. 2D.**
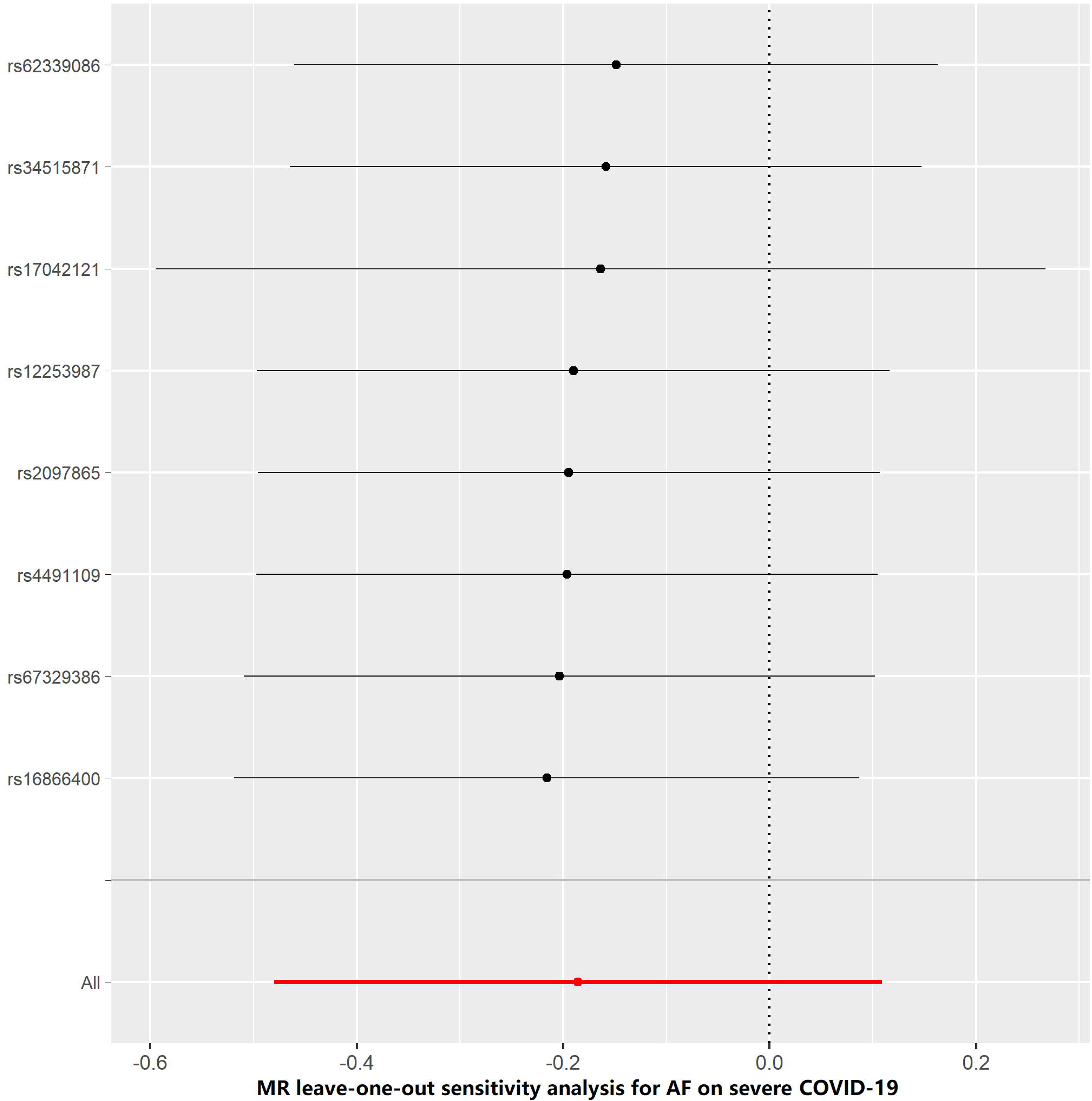
MR leave-one-out analysis for AF on severe COVID-19. Each row represents a MR analysis of AF on severe COVID-19 using all instruments expect for the SNP listed on the y-axis. The point represents the odds ratio with that SNP removed and the line represents 95% confidence interval. COVID-19: Coronavirus disease 2019; AF: Atrial fibrillation; MR: mendelian randomization; SNP, single nucleotide polymorphism

**Figure. 3.**
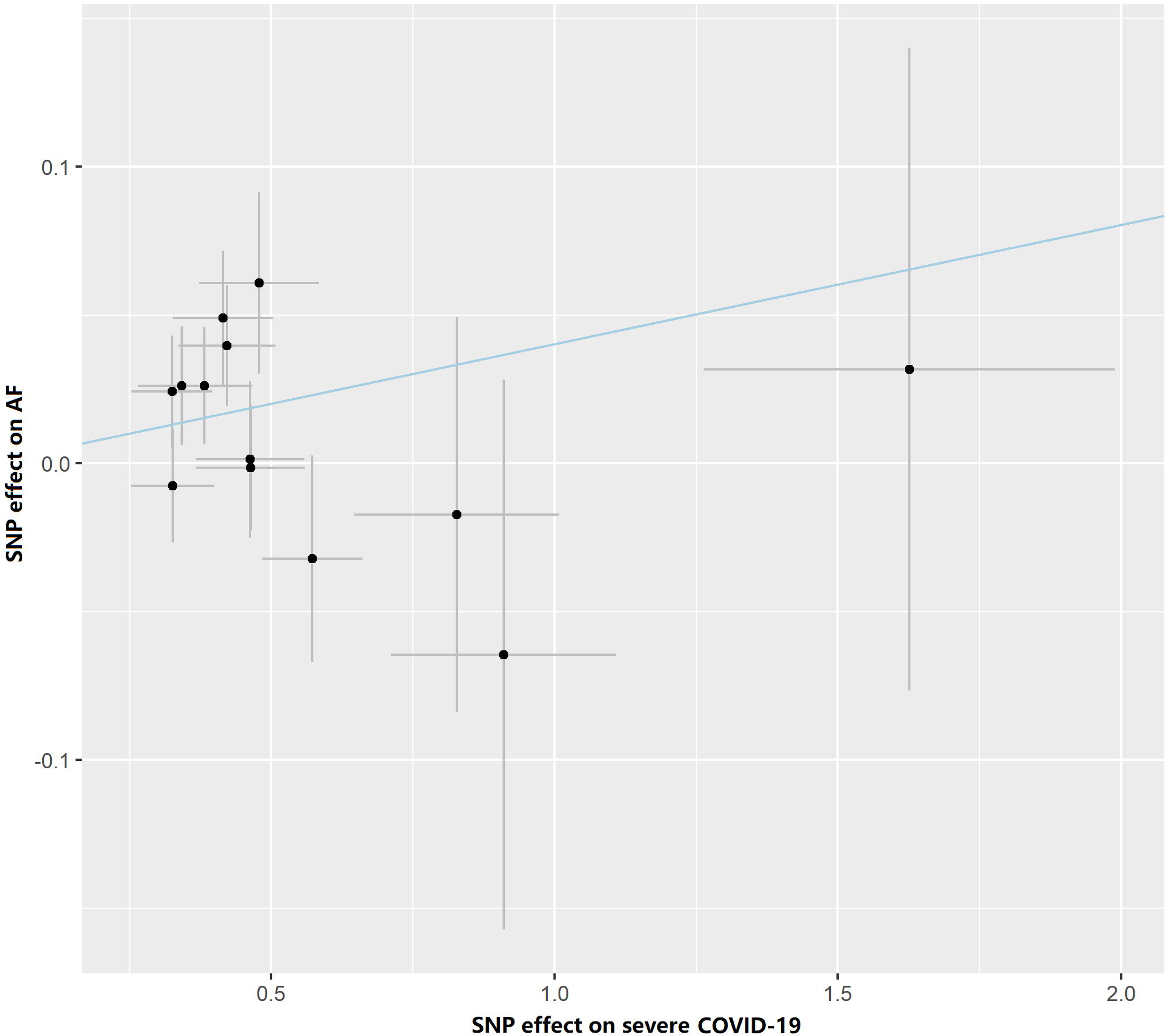
Associations of severe COVID-19 variants with risk of AF performed by inverse-variance weighted method. Circles indicate marginal genetic associations with severe COVID-19 and risk of AF for each variant. Error bars indicate 95% CIs. COVID-19: Coronavirus disease 2019; AF: Atrial fibrillation; MR: mendelian randomization; SNP, single nucleotide polymorphism

## Discussion

To our knowledge, this is the first study to investigate the causal relationship between COVID-19 and AF using a bidirectional MR. Our study showed that severe COVID-19 may causally affect AF through independent pathway.

Our study found that severe COVID-19 was causally associated with the risk of AF, indicating that the incidence of AF in patients with COVID-19 corresponds to the severity of illness. The finding was supported by the observational epidemiology study, which found that cardiac arrests and arrhythmias were more likely to occur in the more severe patients^3^. A worldwide cross-sectional study researched the cardiac arrhythmic in hospitalized COVID-19 patients including severe COVID-19 found that atrial fibrillation was the most common cardiac arrhythmia noted in these patients^15^. Arrhythmia included atrial fibrillation was observed in 7% of patients who did not necessitate ICU treatment and in 44% of subjects who were admitted to an ICU^16^. A multihospital retrospective cohort study of nearly 3,000 patients demonstrated that myocardial injury (MI) is common among patients hospitalized with COVID-19 ^17^, and this may cause AF occurrence^18^.The increasing MR studies have found that coronary artery disease, higher body mass index, lifetime smoking and dyslipidemia were causally associated with the susceptibility of severe COVID-19 ^19-22^, which were also risk factors for AF^23-26^.

We found a possible causal effect of severe COVID-19 on increased risk of AF. Not surprisingly, the severe COVID-19 mainly endanger the elderly and have underlying comorbidities involving diabetes mellitus, hypertension, and coronary heart disease^27 28^, predisposing to AF^29 30^. Plasma angiotensin converting enzyme 2 (ACE2) activity is increased in patients with cardiovascular disease states^31 32^. Previous studies also found that ACE 2 activity may be related to AF^33 34^. Yan et al found that the SARS-CoV-2 has been proved to infect patients through the ACE 2 receptor^35^. Although pulmonary manifestations are its most common clinical symptoms, COVID-19 can lead to systemic inflammation and has varying presentations of cardiac involvement^36^. Severe hypoxic lung disease from severe COVID-19 can trigger atrial arrhythmias. Moreover, the patients may be susceptible to AF due to severe COVID-19-related electrolyte disturbances symptom such as hypokalemia and hyponatremia^37-39^. In addition, the reasons underlying AF also may relate to the proinflammatory effect associated with severe COVID-19. An exaggerated inflammatory response appears a major driver of immunopathology in COVID-19 patients^40^, with benefits of targeting the acute inflammatory response in severe COVID-19 cases^41^. It has been reported that the presence of systemic inflammation determined by elevations in C-reactive protein (CRP) remained a significant predictor of AF^42^, and AF may persist due to structural changes in the atria that are promoted by inflammation^37^.

In our analysis, we did not find any associations between AF and COVID-19 or severe COVID-19. However, it is not clear whether AF would contribute to increasing the risk for severe forms of COVID-19, worse prognosis, or even higher mortality.

The limitations of the current study should be addressed. First, due to the limitation of data resource, stratified analyses or analyses adjusted for other covariates were impossible. In addition, it is hard to represent the universal conclusions for other ethnic groups based on European population.

In conclusion, our study showed that AF might be a consequence rather than a risk factor of severe COVID-19, indicating that the future study should pay attention to the prognosis of severe COVID-19.

## Data Availability

All data generated or analyzed during this study are included in this published article and its supplementary information files.

## Disclosure of Potential Conflicts of Interest

All authors have approved the manuscript and its submission. No potential conflicts of interest were disclosed by the authors.

## Funding/Support

The study was supported by grants from the China-Australian Collaborative Grant (NSFC 81561128020-NHMRC APP1112767).

## Acknowledgements

We want to acknowledge the participants and investigators of the FinnGen study. We thank the COVID-19 host genetics initiative in UK Biobank and severe COVID-19 GWAS Group investigators for making the summary GWAS results publicly accessible.

## Role of the Funder/Sponsor

The funding organization had no role in the design and conduct of the study; collection, management, analysis, and interpretation of the data; preparation, review, or approval of the manuscript; or decision to submit the manuscript for publication.

## Notes

### Competing Interest Statement

The authors have declared no competing interest.

### Clinical Trial

We use Genome-wide association studies (GWAS) data from online.

